# Investigating direct and indirect genetic effects in attention deficit hyperactivity disorder (ADHD) using parent-offspring trios

**DOI:** 10.1101/2022.04.11.22273180

**Authors:** Joanna Martin, Matthew Wray, Sharifah Shameem Agha, Katie J. S. Lewis, Richard J. L. Anney, Michael C. O’Donovan, Anita Thapar, Kate Langley

## Abstract

**Background:** Attention deficit hyperactivity disorder (ADHD) is highly heritable, but little is known about the relative effects of transmitted (i.e. direct) and non-transmitted (i.e. indirect) common variant risks. Using parent-offspring trios, we tested whether polygenic liability for neurodevelopmental and psychiatric disorders and lower cognitive ability is over-transmitted to ADHD probands. We also tested for indirect or ‘genetic nurture’ effects, by examining whether non-transmitted ADHD polygenic liability is elevated. Finally, we examined whether complete trios are representative.

**Methods:** Polygenic risk scores (PRS) for ADHD, anxiety, autism, bipolar disorder, depression, obsessive-compulsive disorder (OCD), schizophrenia, Tourette’s syndrome, and cognitive ability were calculated in UK controls (N=5,081), UK probands with ADHD (N=857), and, where available, their biological parents (N=328 trios), and also a replication sample of 844 ADHD trios.

**Results:** ADHD PRS were over-transmitted and cognitive ability and OCD PRS were under-transmitted to probands. These results were independently replicated. Over-transmission of polygenic liability was not observed for other disorders. Non-transmitted alleles were not enriched for ADHD liability compared to controls. Probands from incomplete trios had more hyperactive-impulsive and conduct disorder symptoms, lower IQ, and lower socioeconomic status than complete trios. PRS did not vary by trio status.

**Conclusions:** The results support direct transmission of polygenic liability for ADHD and cognitive ability from parents to offspring, but not for other neurodevelopmental/psychiatric disorders. They also suggest that non-transmitted neurodevelopmental/psychiatric parental alleles do not contribute indirectly to ADHD via genetic nurture. Furthermore, ascertainment of complete ADHD trios may be non-random, in terms of demographic and clinical factors.

## Introduction

Attention deficit hyperactivity disorder (ADHD) is a highly heritable neurodevelopmental disorder, with robustly associated common genetic risk variants (1). It shares genetic liability with many other neurodevelopmental and psychiatric disorders, as well as lower cognitive ability (1–3). Parents of children with ADHD have a higher prevalence of ADHD and other neurodevelopmental and psychiatric disorders, compared to the general population (4,5). Given that ADHD is highly heritable, cross-generational transmission is likely to be explained by genetic, rather than environmental, factors. However, parents provide the pre- and post-natal environment for their children, both of which impact on early development. It is well established that many environmental exposures are influenced by parental genotypes; known as gene-environment correlation (6). As such, it is possible that ADHD is influenced by parental genetic liability that is not transmitted to the child, in addition to transmitted risks.

ADHD genetic studies frequently use a case-control design (1), but an alternative parent-offspring trio design (using data from a proband with ADHD and both their biological parents) is more suitable for certain research purposes. For example, parent-offspring trios can be used to identify inherited and non-inherited genetic risk variants (7). This design circumvents limitations of the case-control design, such as imperfect case-control matching on confounders (e.g. ancestry) and bias from the use of screened controls in cross-disorder genetic correlation estimates (8). More recently, the trio design has been extended to allow the study of transmission of total common variant liability from parents to offspring (9), as well as to test the impact of indirect genetic effects or ‘genetic nurture’ on children, by examining the contribution of non-transmitted risk alleles (10). Enriched ADHD polygenic liability in non-transmitted parental alleles could exert an effect on the proband through such an indirect ‘genetic nurture’ path.

The first aim of this study was to test whether polygenic risk scores (PRS, or the sum of each individual’s common variant liability) for ADHD, other neurodevelopmental disorders, psychiatric disorders, and lower cognitive ability, are over-transmitted from parents to children with ADHD (i.e. direct genetic effects). Given the high heritability of ADHD, we expect to observe over-transmission of ADHD risk, as well as risk alleles for phenotypes with which ADHD shares genetic liability. This can be tested using the polygenic transmission disequilibrium test (pTDT), which compares proband PRS to the mean of their parents’ PRS (i.e. the common variant liability expected in the proband by chance) (9). Under the hypothesis that manifestation of ADHD depends on direct genetic effects, risk alleles must be transmitted to the proband more often than expected by chance, resulting in a proband PRS greater than the parental mean. When testing for shared cross-disorder genetic effects, this is a more stringent test than case-control analysis, given the limitations of case-control samples outlined above.

The second aim was to investigate whether non-transmitted ADHD risk alleles are elevated in parents of children with ADHD, compared to population controls. Genome-wide association studies (GWAS) of parent-offspring trios use “pseudo-control” alleles, which represent the parental alleles that are not transmitted to the offspring. If the parents themselves have ADHD or have multiple children with ADHD, these non-transmitted alleles could be enriched for ADHD liability compared to the general population, which can potentially reduce genomic discovery power (11,12). Of even greater interest, non-transmitted alleles can exert indirect genetic effects on offspring phenotype and so an enrichment of ADHD risk in non-transmitted alleles compared to controls would be consistent with indirect genetic effects. Such indirect effects may also exist for phenotypes that share genetic liability with ADHD (e.g. other neurodevelopmental and psychiatric disorders, and lower cognitive ability).

A final question is whether children with ADHD recruited into a trio design are representative of the wider clinical ADHD population. The need to obtain DNA from all three individuals in a parent-offspring trio could result in biased ascertainment. This can be a challenge because missing genetic information is unlikely to be missing at random (13). Many children with ADHD do not live with both biological parents, with evidence that ADHD severity is linked to likelihood of a child being in a ‘non-intact’ family (14,15). Previous research by our group suggests that where fathers do not live with the family, or decline to take part in research, children are more likely to have the more severe DSM-IV combined subtype of ADHD and comorbid conduct disorder, compared to those from intact families (16). This could mean that probands from incomplete trios have higher genetic liability for ADHD (and related disorders), affecting the generalisability of studies ascertaining only trios, but this requires investigation.

In this study, we tested the following hypotheses using a UK clinical sample of children diagnosed with ADHD and their biological parents: a) children with ADHD disproportionately inherit liability for neurodevelopmental and psychiatric disorders and lower cognitive ability, b) non-transmitted ADHD polygenic liability is elevated compared to controls (i.e. evidence of genetic nurture), and c) children from incomplete trios have a more severe clinical profile and a higher neurodevelopmental and psychiatric polygenic liability, compared to those from complete trios.

## Methods

### Sample description

Children and young people with ADHD (aged 5–18 years; from hereon: probands) were recruited through Child and Adolescent Psychiatry or Paediatric out-patient clinics across Wales and England. Exclusion criteria were a clinical diagnosis of schizophrenia, history of epilepsy, brain damage or known neurological or genetic disorder. Inclusion criteria were a DSM-III-R or DSM-IV research-based diagnosis for ADHD, confirmed using the Child and Adolescent Psychiatric Assessment (CAPA) (17), a semi-structured diagnostic interview undertaken with parents by trained and supervised psychologists, which assesses DSM-IV inattentive and hyperactive-impulsive symptoms, two additional DSM-III-R symptoms, and impairment. Symptom pervasiveness across settings was confirmed using teacher reports (Child ADHD Teacher Telephone Interview (ChATTI) (18), or Conners’ Teacher Rating Scale (19)).

Written informed consent was obtained from all parents and young people aged 16–18 years old and assent was gained from probands <16 years old. Study approval was obtained from the Northwest England and Wales Multicentre Research Ethics Committees.

ADHD (inattentive and hyperactive-impulsive) symptom scores were generated using DSM-IV criteria. Impairment was assessed using eight items (home life, social interactions, community activities, school, sports/clubs, taking care of oneself, recreational activities, and handling responsibilities). Impairment occurring ‘sometimes’ or ‘often’ was coded as 1 and ‘never’ or ‘rarely’ coded as 0, and items were summed.

The CAPA was also used to assess comorbid symptoms in the preceding 3 months, according to DSM-IV symptom criteria, including conduct disorder (CD), oppositional defiant disorder (ODD), anxiety, and depression. Probands aged 12 and older also completed the child version of the CAPA. A symptom was considered as present if either the parent or proband reported it. Total symptom scores for CD (9 items), ODD (8 items), anxiety (12 items), and depression (8 items) were generated. Autistic traits were assessed using the parent-rated Social Communication Questionnaire (SCQ; 39 items) (20). Full-scale IQ was assessed using the Wechsler Intelligence Scale for Children (WISC), version III or IV, using 10 subtests (21,22). Probands with IQ<70 were considered to have intellectual disability (ID).

Socioeconomic variables (family annual income, parental educational attainment, and parental employment status) and family history of psychiatric disorders were assessed by parental questionnaire. Low income was defined as self-reported gross annual family income <£20,000 (equivalent ∼US$32,000). Parental low educational attainment was defined as parents having left school without qualifications (GCSE or equivalent) at age 16. Socioeconomic status (SES) was classified by the occupation of the main family wage earner using the UK Standard Occupation Classification (23). Two SES categories were defined (low: unskilled workers/unemployed; medium/high: manual and non-manual skilled/partially skilled workers and professional and managerial workers). Family history was based on reported information about first degree relatives (i.e. biological parents and full siblings). Three binary variables were derived, relating to the presence of ADHD, other neurodevelopmental problems (e.g. learning difficulties, dyslexia, dyspraxia), and broadly defined major psychiatric disorders (e.g. depression, bipolar disorder, schizophrenia).

### Genetic data

A detailed description of the genetic data can be found in the **Supplemental Text**. In brief, DNA samples were collected from probands and parents and genotyped, followed by rigorous quality control procedures. Parent-offspring relationships were confirmed using identity-by-descent (IBD) analysis in PLINK. The study ascertained families of European ancestry and ancestry was confirmed using principal components analysis (PCA). For complete parent-offspring trios, non-transmitted parental alleles were extracted using PLINK (using the function --tucc).

PRS were calculated using common autosomal variants based on the following 9 large discovery GWAS of primarily European ancestry: ADHD (1), anxiety disorders (24), autism spectrum disorder (ASD) (25), bipolar disorder (BD) (26), major depressive disorder (MDD) (27), schizophrenia (28), obsessive-compulsive disorder (OCD) (29), Tourette’s syndrome (TS) (30), and cognitive ability (31).

Comparison individuals consisted of 5,081 individuals from the Wellcome Trust Case-Control Consortium (a UK control population sample) not screened for ADHD or other psychiatric disorders (32). The sample has been used previously as controls for a GWAS including a subset of the current ADHD cases (1,33). The ADHD sample (including the ADHD non-transmitted parental pseudo genotypes) was merged with the population controls using shared single nucleotide polymorphisms (SNPs).

The discovery GWAS used to calculate PRS had no overlap with the target ADHD sample. For the merged ADHD-control sample, we obtained GWAS excluding the controls, where possible (all except for MDD and BD).

PRS were calculated using LD-clumping in PLINK (34) for 7 p-value thresholds and the first principal component was extracted and analysed for each discovery phenotype, following the PRS-PCA method, an approach that reduces overfitting, maintaining good power (35); see details in **Supplemental Text**. PRS in the merged ADHD-control sample were standardised using the control population mean and standard deviation. Otherwise, PRS were standardised as z-scores.

PCAiR (36), a package that robustly estimates population structure while taking into account kinship information, was used to extract the top 10 principal components.

### Definition of trio status

857 probands with ADHD (mean age=10.4 years, SD=2.8; N=119 [13.9%] female) from 825 families met inclusion criteria and passed genetic quality control. They were grouped depending on parent-offspring trio status. Complete trios (coded as ‘0’) were defined as families where both parents provided a DNA sample and were confirmed as the biological parents, regardless of whether both parents passed subsequent QC. Incomplete trios (coded as ‘1’) were families where one or both parents did not provide a DNA sample. In families where both parents had provided DNA but one or both parents could not be genotyped due to low sample quality, the probands were unclassified, because parent-offspring relatedness could not be confirmed (N=36). This resulted in a sample of 367 probands from complete trios and 454 probands from incomplete trios.

Additional filters were applied for the analyses of transmitted (pTDT) and non-transmitted alleles, as follows: probands were excluded if parental samples did not pass QC, or if the whole trio was not genotyped on the same array. Only the oldest proband was included for families with multiple probands genotyped (N=39 excluded). This resulted in a sample of 328 trios meeting inclusion criteria for the pTDT and the analysis of non-transmitted parental alleles.

### Analyses

We tested for over-transmission of liability to ADHD, ASD, anxiety, BD, MDD, OCD, schizophrenia, and TS, and under-transmission of liability to cognitive ability, in complete trios using the pTDT (9). This analysis tests whether, on average, the proband PRS deviates significantly from the parental mid-point PRS and is robust to population stratification and other potential confounders (e.g. socioeconomic status).

Next, we compared the ADHD PRS in non-transmitted alleles to population controls. We also explored whether the PRS for other neurodevelopmental and psychiatric disorders and cognitive ability differed between non-transmitted alleles and population controls.

Finally, we compared probands from complete and incomplete trios, in terms of demographic variables, clinical symptoms, family environment and socioeconomic variables, as well as the proband and mother’s PRS for ADHD, other disorders and cognitive ability. Father’s PRS could not be compared as there were only 14 fathers in incomplete trios.

The top 10 ancestry-based principal components were residualised out of the PRS prior to analysis (except for the pTDT). In the analyses comparing probands and mothers based on trio status, genotyping batch was also included as a covariate and we acc0unted for the presence of siblings by specifying family clusters and applying a sandwich estimator to estimate cluster-robust standard errors of regression coefficients. All analyses used generalised estimating equations implemented in the *drgee* package in R. False discovery rate (FDR) correction for multiple testing was applied for the genetic analyses in the primary sample.

### Replication analysis

Independent data from the International Multicentre ADHD Genetics (IMAGE) study (37) were used for replication. The sample consisted of 844 complete parent-offspring trios of probands diagnosed with ADHD (mean age=10.9 years, SD=2.8; N=111 (13.2%) females). Only 616 complete trios matched the WTCCC control population sample ancestry for the analyses of non-transmitted parental alleles. See the **Supplemental Text** for details.

## Results

### Polygenic transmission

PRS for ADHD were over-transmitted [mean(SE)=0.30(0.06)] to probands. There was no evidence of over-transmission of risk alleles for other disorders (Figure 1a & Table S1). Polygenic liabilities for cognitive ability [mean(SE)=-0.33(0.05)] and OCD [mean(SE)=-0.18(0.05)] were under-transmitted. The cognitive ability results were not influenced by comorbid ID; after excluding 28 ADHD probands with ID (IQ<70), the results remained the same [mean(SE)=-0.33(0.05)].

**Figure 1:**
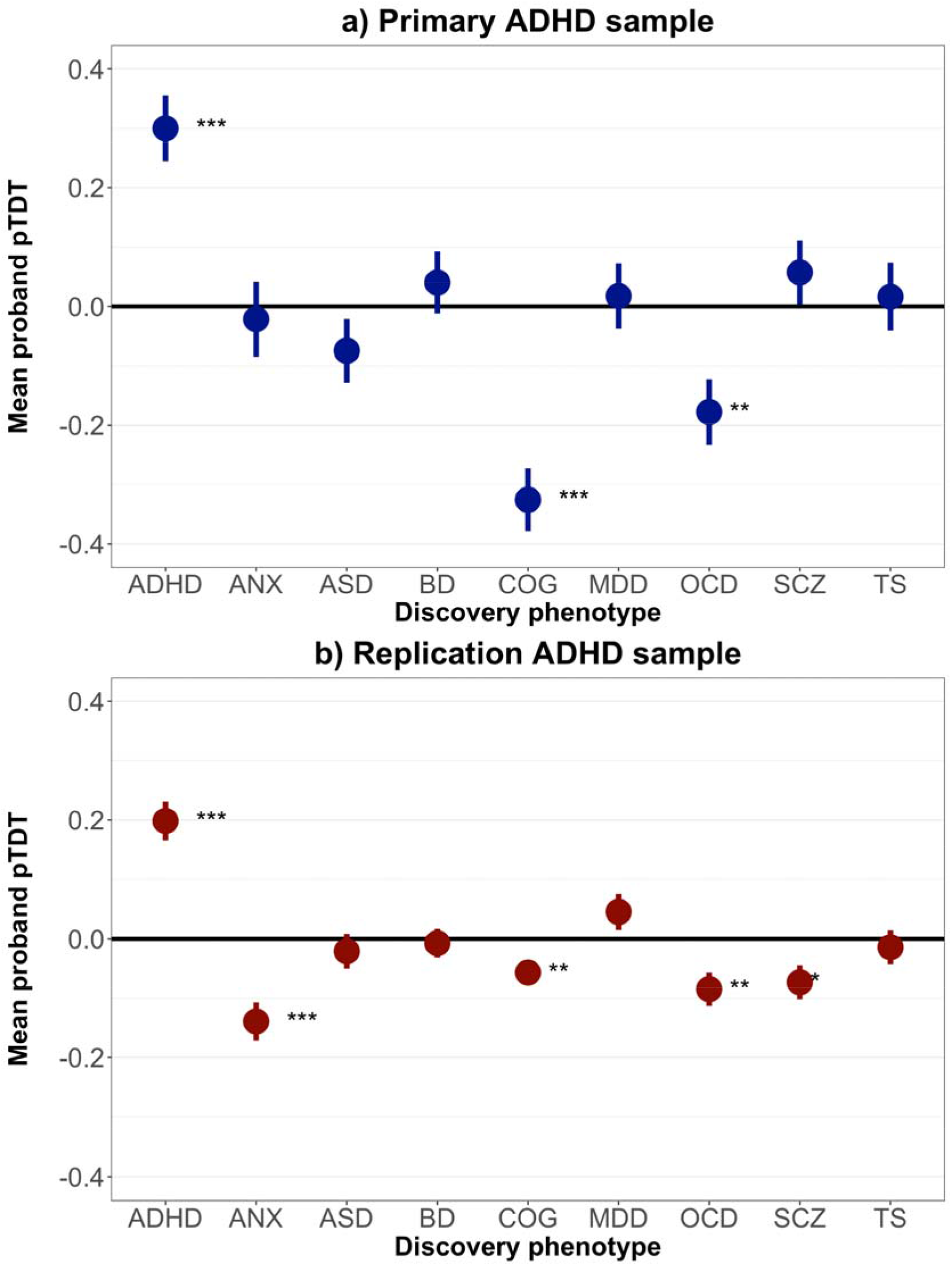
Mean deviation of proband polygenic risk scores from the mid-parent distribution (i.e. standard deviations away from the mid-parent distribution) in ADHD trios, using the a) primary sample (N=328 trios) and the b) replication sample (N=844 trios). ADHD: attention deficit hyperactivity disorder; ANX: anxiety disorders; ASD: autism spectrum disorder; BD: bipolar disorder; COG: cognitive ability; MDD: major depressive disorder; OCD: obsessive-compulsive disorder; pTDT: polygenic transmission disequilibrium test; SCZ: schizophrenia; TS: Tourette’s syndrome. P-values indicate the probability that the mean of the pTDT deviation distribution is 0 (two-sided, one-sample t test). Error bars indicate standard errors. * p<0.05; ** p<0.01; *** p<0.001. P-values shown are corrected for multiple tests for primary analyses and raw p-values are shown for the replication analyses. See Table S1 for detailed results.

The results were independently replicated for ADHD [mean(SE)=0.20(0.03)], cognitive ability [mean(SE)=-0.06(0.02)], and OCD [mean(SE)=-0.08(0.03)] PRS (see Figure 1b & Table S1). Analyses in the replication sample also indicated under-transmission of polygenic liability for anxiety and schizophrenia, but this was not supported by the primary analyses.

### Non-transmitted parental alleles

Figure 2a displays the mean PRS for the 328 complete ADHD trios, separately for probands, mothers, fathers, and non-transmitted parental alleles, relative to the control population sample; see Table S2 for detailed results. There was no evidence supporting elevated ADHD PRS for non-transmitted parental alleles compared to the control population. Exploratory analyses also found little support for elevated non-transmitted parental PRS for other disorders or lower cognitive ability PRS. PRS of probands, fathers, and mothers were elevated for ADHD and lower for cognitive ability, compared to controls. No significant differences were observed for other disorder PRS.

**Figure 2:**
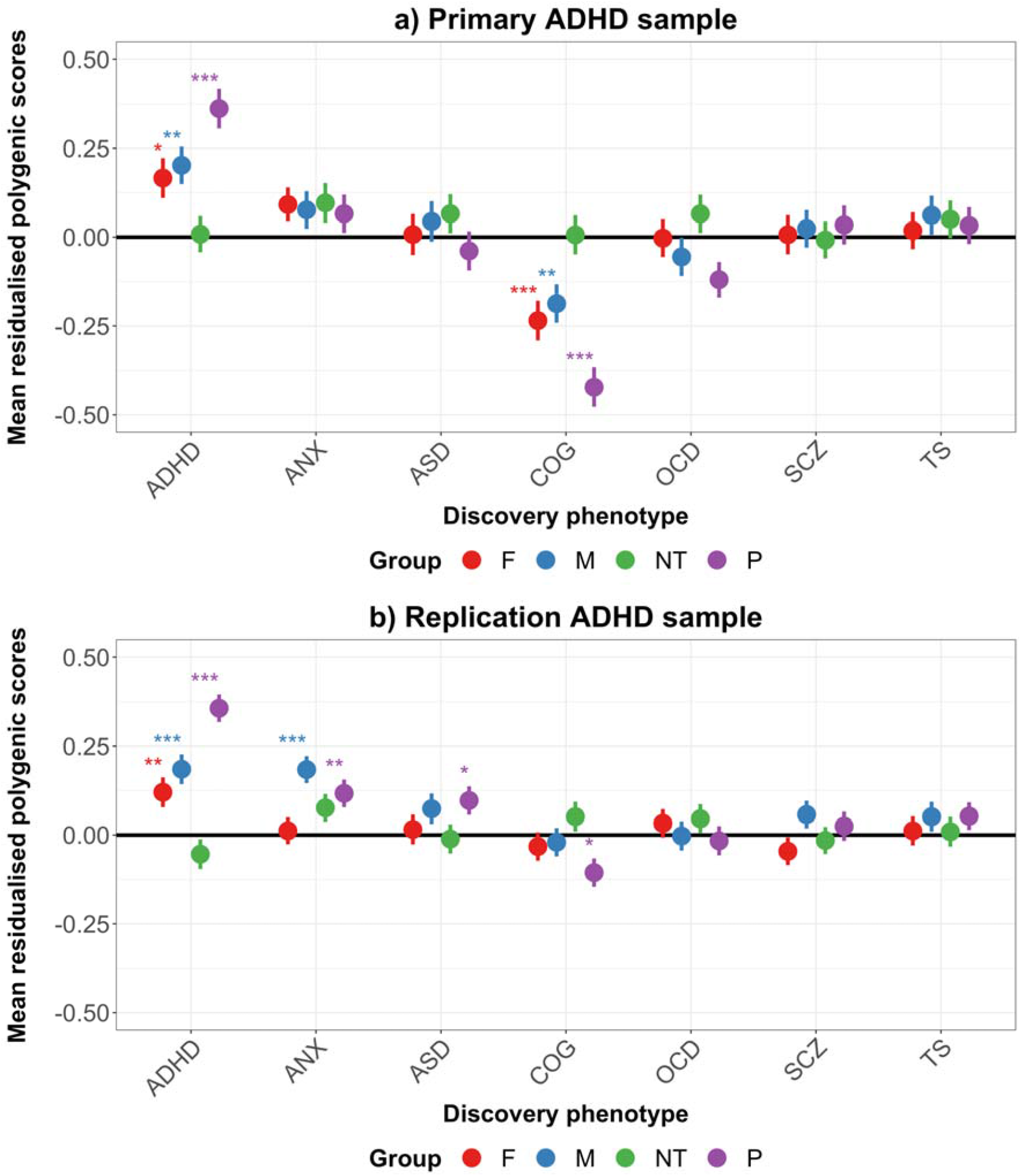
Mean polygenic risk scores in ADHD complete trios: a) primary sample (N=328) and b) replication sample (N=616), in proband (P), fathers (F), mothers (M), and non-transmitted parental alleles (NT), relative to the control population sample (bold horizontal line at y=0). ADHD: attention deficit hyperactivity disorder; ANX: anxiety disorders; ASD: autism spectrum disorder; COG: cognitive ability; SCZ: schizophrenia; OCD: obsessive-compulsive disorder; TS: Tourette’s syndrome. Note: Bipolar disorder and major depressive disorder could not be examined due to inclusion of the control population sample in the discovery genetic studies for those disorders. Error bars indicate standard errors. * p<0.05; ** p<0.01; *** p<0.001. P-values shown are corrected for multiple tests for primary analyses and raw p-values are shown for the replication analyses. See Table S2 for detailed results.

Analyses in the replication sample are shown in Figure 2b and Table S2. The results of the primary analysis were replicated for ADHD PRS. Cognitive ability PRS were lower and ASD PRS were elevated in probands compared to controls. Proband and mothers’ anxiety PRS were elevated compared to controls. There was little evidence of elevated PRS for non-transmitted parental alleles for any phenotypes compared to the control sample and no other group differences were observed.

### Complete and incomplete trios

Analysis of 821 probands (complete trios: N=367, incomplete trios: N=454) indicated that those from incomplete trios were older, had more hyperactive-impulsive ADHD symptoms, lower IQ, more conduct disorder symptoms and were more likely to meet diagnostic criteria for comorbid conduct disorder (see Table 1). Other variables were similar between the groups. Parents of probands from incomplete trios had lower educational attainment, lower annual family income and lower socioeconomic status based on occupation. There were no group differences in family history of ADHD or other disorders.

**Table 1:** Comparison of probands from complete (N=367) and incomplete (N=454) trios in terms of demographic, clinical, socioeconomic, and family history variables.

| Phenotype | Mean (SE)<br>incomplete trios | Mean (SE)<br>complete trios | OR (95% CIs)** | p |
| --- | --- | --- | --- | --- |
| Proband age* | 10.70 (0.13) | 10.10 (0.15) | 1.07 (1.02-1.13) | **6.3 × 10<sup>-3</sup>** |
| Inattentive symptoms | 7.48 (0.08) | 7.27 (0.09) | 1.07 (0.98-1.17) | 0.11 |
| Hyperactive-impulsive symptoms | 7.82 (0.07) | 7.65 (0.08) | 1.12 (1.02-1.24) | **0.019** |
| ADHD impairment | 6.88 (0.07) | 6.69 (0.09) | 1.11 (0.99-1.24) | 0.087 |
| IQ | 82.90 (0.67) | 85.50 (0.71) | 0.99 (0.98-1.00) | **0.041** |
| Autistic traits | 13.50 (0.36) | 12.80 (0.46) | 1.02 (0.99-1.04) | 0.23 |
| Anxiety symptoms | 1.02 (0.09) | 0.93 (0.10) | 1.03 (0.93-1.13) | 0.60 |
| Depressive symptoms | 1.42 (0.07) | 1.21 (0.07) | 1.09 (0.98-1.22) | 0.11 |
| ODD symptoms | 4.11 (0.11) | 3.90 (0.13) | 1.04 (0.98-1.10) | 0.19 |
| CD symptoms | 1.53 (0.08) | 1.16 (0.09) | 1.12 (1.02-1.23) | **0.012** |
| Phenotype | N (%)<br>incomplete trios | N (%)<br>complete trios | OR (95% CIs) | p |
| Proband sex (male) | 72 (15.9) | 46 (12.5) | 1.35 (0.91-2.01) | 0.13 |
| Low family income | 212 (72.4) | 105 (50.5) | 2.64 (1.78-3.90) | **1.3 × 10<sup>-6</sup>** |
| Low parental education | 100 (31.6) | 47 (21.9) | 1.67 (1.10-2.52) | **0.015** |
| Low family SES | 230 (59.9) | 125 (39.1) | 2.35 (1.71-3.22) | **1.1 × 10<sup>-7</sup>** |
| Intellectual disability | 58 (13.6) | 29 (8.2) | 1.27 (1.00-1.61) | 0.055 |
| CD diagnosis | 107 (23.7) | 60 (16.4) | 1.51 (1.05-2.18) | **0.025** |
| Family history: ADHD | 75 (23.7) | 60 (25.0) | 0.92 (0.60-1.42) | 0.71 |
| Family history: Other NDs | 40 (12.9) | 32 (13.4) | 0.95 (0.56-1.60) | 0.84 |
| Family history: Major psychiatric disorders | 139 (42.4) | 89 (37.4) | 1.24 (0.87-1.77) | 0.24 |

\* Proband age is included as a covariate in all other analyses. \*\* Probands from complete trios were coded as '0' and those from incomplete trios were coded as '1', therefore OR>1 can be interpreted as an increase of the variable in the probands from incomplete trios. ODD: oppositional defiant disorder; CD: conduct disorder; SES: socioeconomic status; ND: neurodevelopmental disorders.

Analysis of probands’ and mothers’ PRS indicated several nominally significant effects (see Table 2). Probands from incomplete trios had higher PRS for BD and OCD, and higher maternal PRS for ADHD. On the other hand, mothers’ PRS for schizophrenia were lower in incomplete trios. However, these effects did not withstand multiple testing correction.

**Table 2:**
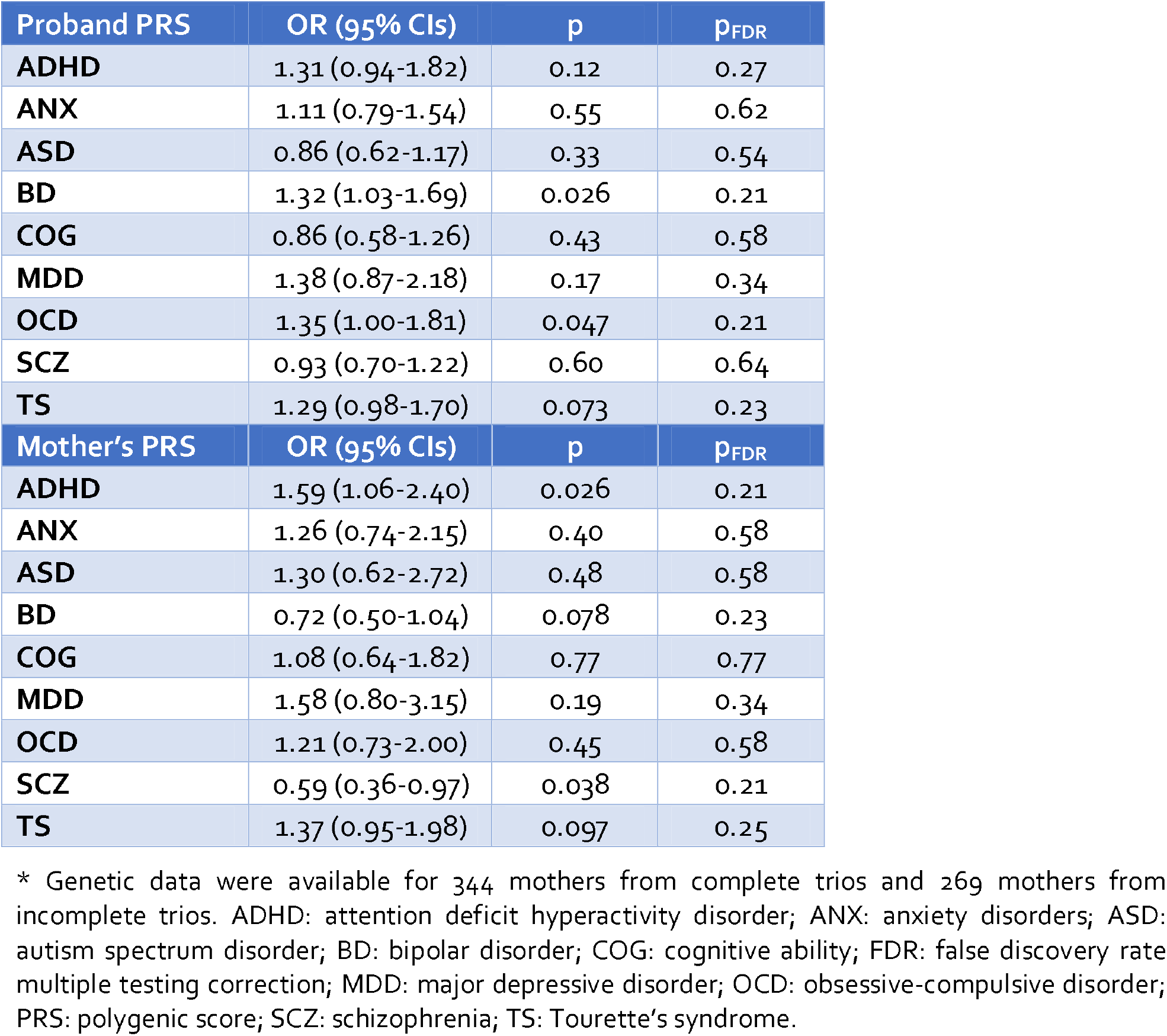
Comparison of polygenic risk scores in probands and mothers from complete (N=367)* and incomplete (N=454) trios.

| Proband PRS | OR (95% CIs) | p | p<sub>FDR</sub> |
| --- | --- | --- | --- |
| ADHD | 1.31 (0.94-1.82) | 0.12 | 0.27 |
| ANX | 1.11 (0.79-1.54) | 0.55 | 0.62 |
| ASD | 0.86 (0.62-1.17) | 0.33 | 0.54 |
| BD | 1.32 (1.03-1.69) | 0.026 | 0.21 |
| COG | 0.86 (0.58-1.26) | 0.43 | 0.58 |
| MDD | 1.38 (0.87-2.18) | 0.17 | 0.34 |
| OCD | 1.35 (1.00-1.81) | 0.047 | 0.21 |
| SCZ | 0.93 (0.70-1.22) | 0.60 | 0.64 |
| TS | 1.29 (0.98-1.70) | 0.073 | 0.23 |
| Mother's PRS | OR (95% CIs) | p | p<sub>FDR</sub> |
| ADHD | 1.59 (1.06-2.40) | 0.026 | 0.21 |
| ANX | 1.26 (0.74-2.15) | 0.40 | 0.58 |
| ASD | 1.30 (0.62-2.72) | 0.48 | 0.58 |
| BD | 0.72 (0.50-1.04) | 0.078 | 0.23 |
| COG | 1.08 (0.64-1.82) | 0.77 | 0.77 |
| MDD | 1.58 (0.80-3.15) | 0.19 | 0.34 |
| OCD | 1.21 (0.73-2.00) | 0.45 | 0.58 |
| SCZ | 0.59 (0.36-0.97) | 0.038 | 0.21 |
| TS | 1.37 (0.95-1.98) | 0.097 | 0.25 |

\* Genetic data were available for 344 mothers from complete trios and 269 mothers from incomplete trios. ADHD: attention deficit hyperactivity disorder; ANX: anxiety disorders; ASD: autism spectrum disorder; BD: bipolar disorder; COG: cognitive ability; FDR: false discovery rate multiple testing correction; MDD: major depressive disorder; OCD: obsessive-compulsive disorder; PRS: polygenic score; SCZ: schizophrenia; TS: Tourette's syndrome.

## Discussion

We used a parent-offspring trio sample of probands with ADHD to test three hypotheses: over-transmission of polygenic liability from parents to probands, elevated polygenic liability in non-transmitted parental alleles, and non-representativeness of ADHD trios. We found robust evidence of over-transmission of ADHD and lower cognitive ability polygenic liability. This was replicated in an independent ADHD sample and consistent with case-control analysis. Unexpectedly, we found evidence of under-transmission of OCD PRS. Also, we found limited evidence of over-transmission or case-control differences for other disorder PRS. Parental non-transmitted alleles related to ADHD and other phenotypes were not elevated compared to a control population; i.e. we observed no evidence of “genetic nurture”. Finally, we observed several clinical and socioeconomic differences between probands from complete and incomplete trios, but no robust differences in PRS.

Over-transmission of ADHD polygenic liability, while not previously tested in ADHD using the pTDT, was expected. Interestingly, the magnitude of the effect sizes observed for ADHD and cognitive ability were similar in the primary sample, despite differences in size and genetic architecture of these two discovery GWAS. ADHD is strongly associated with lower cognitive ability and twin studies show a genetic correlation of 91% between ADHD and intellectual disability (38). Excluding 28 probands with intellectual disability did not affect the results. These results were replicated in an independent and larger ADHD sample from various European countries, although the effect sizes were smaller in the replication sample, particularly for cognitive ability. However, the primary sample’s IQ is lower than the population average [mean(SE)=84.2(0.48)], as expected for ADHD (39). The mean IQ of the replication sample was comparable to the population average [mean(SE)=100.4(0.66)] and only 10 probands had IQ<70, which could explain the lower effect size observed. However, the replicated result suggests that over-transmission of lower cognitive ability polygenic liability is not entirely explained by individuals with lower IQ.

Previous estimates of genetic correlation between ADHD and cognitive ability (r_g_=-0.41) are of a similar magnitude to those between ADHD and MDD (r_g_=0.42), ASD (r_g_=0.36), and anxiety (r_g_=0.33) (1,24,25,27). Despite these similar genetic correlations, we do not see evidence of over-transmission of PRS or case-control differences for these other disorders in our study. It is possible that genetic correlations estimated in previous studies have been over-estimated, for example by inclusion of comorbid cases in discovery GWAS (e.g. individuals with both ADHD and MDD in GWAS of each disorder) or through use of over-screened controls (8). It is also likely that differences in genetic architecture across phenotypes (e.g. smaller total contribution of common variants to heritability for MDD and other disorders) impacted these results and that larger sample sizes than were available in this study are needed to detect shared genetic effects across ADHD and other disorders. Alternatively, it could be that probands with ADHD who over-inherit polygenic liability for both ADHD and other disorders from their parents may show a different phenotype (e.g. ADHD and comorbid bipolar disorder or psychosis) and meet study exclusion criteria or are less likely to take part in genetic studies and were thus missing from our sample. The replicated under-transmission of OCD PRS also contrasts with previously reported positive genetic correlations (r_g_∼0.15) between ADHD and OCD (40) and needs further consideration in future studies. It is possible that probands with a comorbid presentation of ADHD and OCD were less likely to have been included as trios in the current study. This possibility is supported by the slightly higher observed OCD PRS in the incomplete trios, albeit this result did not survive correction for multiple testing.

Contrary to the second hypothesis we tested, we found no evidence in either sample of elevated polygenic liability for ADHD in the non-transmitted parental alleles, compared to a control population. Similarly, the results of our exploratory analyses indicated no enrichment of non-transmitted parental alleles for polygenic liability for other neurodevelopmental/psychiatric disorders or for lower cognitive ability. These results are consistent with a recent study and also a pre-print, which examined ADHD symptoms in the general population (41,42). Our study is different because it focuses on clinical ADHD diagnosis. These studies found that the non-transmitted parental alleles for ADHD and educational attainment do not contribute to risk of ADHD symptoms in a general childhood population, in contrast to transmitted parental alleles (41,42). Together with our results and the limited role of shared environmental risks in ADHD reported in twin studies (43), this indicates that parental polygenic liability for ADHD primarily impacts on child ADHD risk via direct genetic transmission, rather than indirect genetic “nurture” effects.

Our results also indicate that common variant discovery GWAS of ADHD may not be adversely impacted by the use of pseudo-controls from parent-offspring trios relative to case-control samples, as has been previously suggested (11). It is likely that non-transmitted parental risk alleles could be enriched in a sub-group (e.g. families with multiple affected children) or that our use of an unscreened population sample (making this a conservative test) impacted the results. This needs to be investigated in future studies.

Comparison of polygenic liability in complete and incomplete trios indicated weak differences; probands from incomplete trios had elevated PRS for bipolar disorder and OCD and their mothers had elevated ADHD PRS but decreased schizophrenia PRS. However, these results did not withstand multiple testing correction and require follow-up. We were not able to replicate these analyses as incomplete trios were screened out of the IMAGE cohort. There were no differences in family history of ADHD and other disorders. These results indicate that there are no substantial genetic differences (in terms of PRS or family history) in probands and their mothers depending on whether they were recruited to the study as part of a complete trio or not. As such, ADHD trio samples are reasonably representative of clinical ADHD samples in terms of polygenic background and our results examining polygenic over-transmission and non-transmitted parental alleles reflect typical UK clinical populations.

We observed several demographic and clinical differences depending on trio status. Building on a previous study using a subset of 241 (28%) probands drawn from the current sample (16), we observed that probands from incomplete trios had a more severe clinical profile, with more hyperactive-impulsive ADHD and conduct disorder symptoms as well as lower IQ. The probands from incomplete trios were more likely to meet diagnostic criteria for conduct disorder, as previously reported (16). We found no differences in inattentive symptoms, ADHD impairment, or symptoms of ODD, anxiety or depression. Thus, the group of probands from incomplete trios were not generally more impaired but rather showed specific differences in clinical profile, relating to cognitive ability and behavioural symptoms. We also found differences in socioeconomic variables, which may be explained by the lower socioeconomic status of single-parent families (44), who constitute a subset of the incomplete trios. We defined trio status based on availability of DNA from both biological parents, compared to the previous definition of whether fathers of probands with ADHD live with the family and take part in research (16). Though these definitions will overlap, our study is more specifically relevant to considerations of whether trio samples in genetic studies are representative of an ADHD clinical sample and our results indicate that this is not entirely the case.

We were unable to compare fathers’ polygenic profiles given few fathers in incomplete trios. We were also unable to compare non-transmitted parental alleles for MDD and BD to the control population, owing to the inclusion of the controls in those discovery GWAS. Although we found only weak evidence of differences in polygenic liability in complete and incomplete trios, this may have impacted the analysis of non-transmitted parental alleles, which needs to be studied further.

Overall, our results suggest that probands with ADHD over-inherit not just polygenic liability for ADHD but also for lower cognitive ability. We find no evidence of enrichment of polygenic liability for neurodevelopmental or psychiatric phenotypes in non-transmitted parental alleles, suggesting that genetically influenced nurture (as captured by the PRS we tested) does not contribute to ADHD risk. Finally, our results indicate that probands who are recruited to trio-based genetic study designs may not be entirely representative of clinical samples, showing a somewhat less severe clinical profile and higher family socioeconomic status.

## Data Availability

The primary ADHD proband data used in the present study are available via the Psychiatric Genomics Consortium. Complete parent-offspring trio data will be made available in future, see medRxiv pre-print titled "DRAGON-Data: A platform and protocol for integrating genomic and phenotypic data across large psychiatric cohorts".
The International Multisite ADHD Genetics (IMAGE) data are available via dbGaP.

https://www.med.unc.edu/pgc/shared-methods/open-source-philosophy/

https://www.medrxiv.org/content/10.1101/2022.01.18.22269463v1.full-text

https://www.ncbi.nlm.nih.gov/projects/gap/cgi-bin/study.cgi?study_id=phs000016.v2.p2

## Data Availability

The primary ADHD proband data used in the present study are available via the Psychiatric Genomics Consortium. Complete parent-offspring trio data will be made available in future, see medRxiv pre-print titled "DRAGON-Data: A platform and protocol for integrating genomic and phenotypic data across large psychiatric cohorts". 
The International Multisite ADHD Genetics (IMAGE) data are available via dbGaP.

https://www.med.unc.edu/pgc/shared-methods/open-source-philosophy/

https://www.medrxiv.org/content/10.1101/2022.01.18.22269463v1.full-text

https://www.ncbi.nlm.nih.gov/projects/gap/cgi-bin/study.cgi?study_id=phs000016.v2.p2

## Acknowledgements

The work was supported by funding from the Wellcome Trust (grant no. 079711 & 220488/Z/20/Z), Medical Research Council Centre (grant no. MR/L010305/1), Health and Care Research Wales (grant no. 514032), Action Medical Research and Baily Thomas. We also acknowledge the support of the Supercomputing Wales project, which is part-funded by the European Regional Development Fund (ERDF) via Welsh Government. JM was supported by a NARSAD Young Investigator Grant from the Brain & Behavior Research Foundation (grant no. 27879). This work was supported by the Wolfson Centre for Young People’s Mental Health. This research was funded in whole, or in part, by the Wellcome Trust. For the purpose of open access, the author has applied a CC BY public copyright licence to any Author Accepted Manuscript version arising from this submission.

We acknowledge dbGaP, via the dbGaP accession: phs000016.v2.p2 https://www.ncbi.nlm.nih.gov/projects/gap/cgi-bin/study.cgi?study_id=phs000016.v2.p2. Funding support for the International Multisite ADHD Genetics (IMAGE) project was provided by NIH grants R01MH62873 and R01MH081803 to S.V. Faraone and the genotyping of samples was provided through the Genetic Association Information Network (GAIN). The dataset(s) used for the analyses described in this manuscript were obtained from the database of Genotypes and Phenotypes (dbGaP) found at http://www.ncbi.nlm.nih.gov/gap through dbGaP accession number 26394. Samples and associated phenotype data for the International Multi-Center ADHD Genetics Project were provided by the following investigators: S. Faraone (PI), R. Anney, P. Asherson, J. Sergeant, R. Ebstein, B. Franke, M. Gill, A. Miranda, F. Mulas, R. Oades, H. Roeyers, A. Rothenberger, T. Banaschewski, J. Buitelaar, E. Sonuga-Barke (site PIs), M. Daly, C. Lange, N. Laird, J. Su, and B. Neale (statistical analysis team).

This study makes use of data generated by the Wellcome Trust Case-Control Consortium. A full list of the investigators who contributed to the generation of the data is available from www.wtccc.org..uk. Funding for the project was provided by the Wellcome Trust under award 076113, 085475 and 090355.

## Author contributions

Study conception & design: Joanna Martin, Kate Langley

Data preparation and analysis: Joanna Martin, Matthew Wray, Katie Lewis, Richard Anney

Writing (original draft): Joanna Martin, Matthew Wray

Writing (review & editing): All authors

## References

1. Demontis D, Walters RK, Martin J, Mattheisen M, Als TD, Agerbo E, et al. Discovery of the first genome-wide significant risk loci for attention deficit/hyperactivity disorder. Nat Genet. 2019 Nov 26;51:63–75.

2. Andersson A, Tuvblad C, Chen Q, Du Rietz E, Cortese S, Kuja-Halkola R, et al. Research Review: The strength of the genetic overlap between ADHD and other psychiatric symptoms – a systematic review and meta-analysis. J Child Psychol Psychiatry Allied Discip. 2020;61(11):1173–83.

3. Lee PH, Anttila V, Won H, Feng YCA, Rosenthal J, Zhu Z, et al. Genomic Relationships, Novel Loci, and Pleiotropic Mechanisms across Eight Psychiatric Disorders. Cell. 2019;179(7):1469-1482.e11.

4. Thapar A. Discoveries on the genetics of ADHD in the 21st century: New findings and their implications. Vol. 175, American Journal of Psychiatry. American Psychiatric Association; 2018. p. 943–50.

5. Agha SS, Zammit S, Thapar A, Langley K. Are parental ADHD problems associated with a more severe clinical presentation and greater family adversity in children with ADHD? Eur Child Adolesc Psychiatry. 2013;22(6):369–77.

6. Thapar A, Rice F. Family-Based Designs that Disentangle Inherited Factors from Pre-and Postnatal Environmental Exposures: In Vitro Fertilization, Discordant Sibling Pairs, Maternal versus Paternal Comparisons, and Adoption Designs. Cold Spring Harb Perspect Med. 2021 Mar 1;11(3):a038877.

7. Martin J, Hosking G, Wadon M, Agha SS, Langley K, Rees E, et al. A brief report: de novo copy number variants in children with attention deficit hyperactivity disorder. Transl Psychiatry. 2020 Dec 1;10(1).

8. Kendler KS, Chatzinakos C, Bacanu S. The impact on estimations of genetic correlations by the use of super-normal, unscreened, and family-history screened controls in genome wide case–control studies. Genet Epidemiol. 2020 Apr 21;44(3):283–9.

9. Weiner DJ, Wigdor EM, Ripke S, Walters RK, Kosmicki JA, Grove J, et al. Polygenic transmission disequilibrium confirms that common and rare variation act additively to create risk for autism spectrum disorders. Nat Genet. 2017 May 15;49(7):978–85.

10. Kong A, Thorleifsson G, Frigge ML, Vilhjalmsson BJ, Young AI, Thorgeirsson TE, et al. The nature of nurture: Effects of parental genotypes. Science (80-). 2018 Jan 26;359(6374):424–8.

11. Peyrot WJ, Boomsma DI, Penninx BWJH, Wray NR. Disease and Polygenic Architecture: Avoid Trio Design and Appropriately Account for Unscreened Control Subjects for Common Disease. Am J Hum Genet. 2016 Feb;98(2):382–91.

12. Klei L, Sanders SJ, Murtha MT, Hus V, Lowe JK, Willsey AJ, et al. Common genetic variants, acting additively, are a major source of risk for autism. Mol Autism. 2012 Jan;3(1):9.

13. Martin J, Tilling K, Hubbard L, Stergiakouli E, Thapar A, Davey Smith G, et al. Association of Genetic Risk for Schizophrenia With Nonparticipation Over Time in a Population-Based Cohort Study. Am J Epidemiol. 2016 May 10;183(12):1149–58.

14. Hurtig T, Ebeling H, Taanila A, Miettunen J, Smalley S, McGough J, et al. ADHD and comorbid disorders in relation to family environment and symptom severity. Eur Child Adolesc Psychiatry. 2007 Sep;16(6):362–9.

15. Hurtig T, Taanila A, Ebeling H, Miettunen J, Moilanen I. Attention and behavioural problems of Finnish adolescents may be related to the family environment. Eur Child Adolesc Psychiatry 2005 148. 2005 Dec;14(8):471–8.

16. West A, Langley K, Hamshere ML, Kent L, Craddock N, Owen MJ, et al. Evidence to suggest biased phenotypes in children with Attention Deficit Hyperactivity Disorder from completely ascertained trios. Mol Psychiatry. 2002 Jan;7(9):962–6.

17. Angold A, Prendergast M, Cox A, Harrington R, Simonoff E, Rutter M. The Child and Adolescent Psychiatric Assessment (CAPA). Psychol Med. 1995;25(4):739–53.

18. Holmes J, Lawson D, Langley K, Fitzpatrick H, Trumper A, Pay H, et al. The Child Attention-Deficit Hyperactivity Disorder Teacher Telephone Interview (CHATTI): reliability and validity. Br J Psychiatry. 2004;184:74–8.

19. Conners CK, Sitarenios G, Parker JD, Epstein JN. Revision and restandardization of the Conners Teacher Rating Scale (CTRS-R): factor structure, reliability, and criterion validity. J Abnorm Child Psychol. 1998;26(4):279–91.

20. Rutter M, Bailey A, Lord C, Berument SK. Social communication questionnaire. Los Angeles, CA West Psychol Serv. 2003;

21. Wechsler D. Wechsler Intelligence Scale for Children. 3rd Edition. The Psychological Association; 1992.

22. Wechsler D. Wechsler Intelligence Scale for Children -Fourth Edition (WISC-IV) administration and scoring manual. San Antonio, Tx: The Psychological Association; 2003.

23. Office of National Statistics. Infant feeding survery, 2000. London: The Stationary Office; 2001.

24. Purves KL, Coleman JRI, Meier SM, Rayner C, Davis KAS, Cheesman R, et al. A major role for common genetic variation in anxiety disorders. Mol Psychiatry. 2019 Nov 20;1–12.

25. Grove J, Ripke S, Als TD, Mattheisen M, Walters RK, Won H, et al. Identification of common genetic risk variants for autism spectrum disorder. Nat Genet. 2019 Jan 1;51(3):431–44.

26. Stahl EA, Breen G, Forstner AJ, McQuillin A, Ripke S, Trubetskoy V, et al. Genome-wide association study identifies 30 loci associated with bipolar disorder. Nat Genet. 2019;51(5):793–803.

27. Wray N, Ripke S, Mattheisen M, Traylor Trzaskowski M, Byrne E, Abdellaoui A, et al. Genome-wide association analyses identify 44 risk variants and refine the genetic architecture of major depression. Nat Genet. 2018 May 1;50(5):668–81.

28. Schizophrenia Working Group of the Psychiatric Genomics Consortium. Mapping genomic loci prioritises genes and implicates synaptic biology in schizophrenia. medRxiv. 2020 Sep 13;2020.09.12.20192922.

29. Arnold PD, Askland KD, Barlassina C, Bellodi L, Bienvenu OJ, Black D, et al. Revealing the complex genetic architecture of obsessive-compulsive disorder using meta-analysis. Mol Psychiatry. 2018 May 1;23(5):1181–8.

30. Yu D, Sul JH, Tsetsos F, Nawaz MS, Huang AY, Zelaya I, et al. Interrogating the Genetic Determinants of Tourette’s Syndrome and Other Tic Disorders Through Genome-Wide Association Studies. https://doi.org/101176/appi.ajp201818070857. 2019 Mar 1;176(3):217–27.

31. Savage J, Jansen P, Stringer S, Watanabe K, Bryois J, de Leeuw C, et al. Genome-wide association meta-analysis in 269,867 individuals identifies new genetic and functional links to intelligence. Nat Genet. 2018 Jul 1;50(7):912–9.

32. The Wellcome Trust Case Control Consortium. Genome-wide association study of 14,000 cases of seven common diseases and 3,000 shared controls. Nature. 2007;447(7145):661–78.

33. Stergiakouli E, Hamshere M, Holmans P, Langley K, Zaharieva I, Hawi Z, et al. Investigating the contribution of common genetic variants to the risk and pathogenesis of ADHD. Am J Psychiatry. 2012 Feb;169(2):186–94.

34. Purcell S, Neale B, Todd-Brown K, Thomas L, Ferreira MA, Bender D, et al. PLINK: a tool set for whole-genome association and population-based linkage analyses. Am J Hum Genet. 2007;81(3):559–75.

35. Coombes BJ, Ploner A, Bergen SE, Biernacka JM. A principal component approach to improve association testing with polygenic risk scores. Genet Epidemiol. 2020 Jul 21;44(7):gepi.22339.

36. Conomos MP, Miller MB, Thornton TA. Robust inference of population structure for ancestry prediction and correction of stratification in the presence of relatedness. Genet Epidemiol. 2015;39(4):276–93.

37. Neale BM, Lasky-Su J, Anney R, Franke B, Zhou K, Maller J, et al. Genome-wide association scan of attention deficit hyperactivity disorder. Am J Med Genet Part B Neuropsychiatr Genet. 2008;3255(315):1337–44.

38. Faraone S V., Ghirardi L, Kuja-Halkola R, Lichtenstein P, Larsson H, Faraone SV, et al. The Familial Co-Aggregation of Attention-Deficit/Hyperactivity Disorder and Intellectual Disability: A Register-Based Family Study. J Am Acad Child Adolesc Psychiatry. 2016 Dec;0(0):15020–1266.

39. Kuntsi J, Eley TC, Taylor A, Hughes C, Asherson P, Caspi A, et al. Co-occurrence of ADHD and low IQ has genetic origins. Am J Med Genet Part B Neuropsychiatr Genet. 2004;124(1):41–7.

40. Strom NI, Yu D, Gerring ZF, Halvorsen MW, Abdellaoui A, Rodriguez-Fontenla C, et al. Genome-wide association study identifies newlocus associated with OCD. medRxiv. 2021;

41. de Zeeuw EL, Hottenga JJ, Ouwens KG, Dolan C V., Ehli EA, Davies GE, et al. Intergenerational Transmission of Education and ADHD: Effects of Parental Genotypes. Behav Genet. 2020 Jul 1;50(4):221–32.

42. Pingault J-B, Barkhuizen W, Wang B, Hannigan LJ, Eilertsen EM, Andreassen OA, et al. Identifying intergenerational risk factors for ADHD symptoms using polygenic scores in the Norwegian Mother, Father and Child Cohort. medRxiv. 2021 Feb 20;2021.02.16.21251737.

43. Faraone S V., Larsson H. Genetics of attention deficit hyperactivity disorder. Mol Psychiatry. 2018;1–14.

44. Child Poverty Action Group. Child Poverty Facts and Figures. 2021.

