## Supplemental Materials for "Investigating direct and indirect genetic effects in attention deficit hyperactivity disorder (ADHD) using parent-offspring trios"

**Supplemental Text**

*Quality control of genetic data*

DNA samples (from saliva or blood) were collected from children and where possible, both biological parents. A subset of the children were genotyped using the Illumina Human660W-Quad BeadChip for the purpose of a previous genome-wide association study (1). Children and parents were also genotyped using a customised version of the PsychChip. Quality control (QC) was performed within batch, as follows: SNPs were aligned to the Haplotype Reference Consortium (2) data using GenomeHarmoniser (3), SNPs were removed if they had MAF<0.01, genotyping rate<0.95, or HWE p<10^-6^, and individuals were removed if they had missingness>0.05, sex discrepancy, or were duplicate samples. The data were merged with other samples on the same or equivalent platform and the QC parameters were reapplied, then the samples were imputed using the Michigan Imputation Server (using Eagle v2.4 for phasing, Minimac4 for imputation, and the HRC V1.1 imputation reference panel) (4). After imputation, dosage data were converted to best guess genotype data using PLINK-2.

Post-imputation QC filters were genotype probability >0.9 per individual, missingness <0.03, MAF>0.01, HWE p<10^-4^, and INFO/r^2^>0.8. Batches were merged, including only overlapping SNPs and excluding ambiguous (CG/AT) variants and SNPs with inconsistent alleles. Family relationships were confirmed using identity-by-descent (IBD) and Mendel analyses in PLINK. SNPs with >5 Mendel errors were excluded, and all remaining Mendel errors were set to missing. Samples that were not related as expected based on family structure were excluded.

The study primarily ascertained individuals of European ancestry. PCAiR (5), a package that robustly estimates population structure while taking into account kinship information in the sample, was used to run a principal components analysis (PCA), on an LD-pruned set of common (MAF>0.05) markers. Non-European samples were excluded based on excessive deviation from the sample mean for the top 5 PCs (N=29 samples excluded). GWAS of batch effects were run on unrelated samples and SNPs associated with batch (p<0.01) were excluded (N=104,416 SNPs excluded). Only samples from children and parents who passed all the above QC were included in the study.

Non-transmitted parental alleles were derived as pseudo-controls using PLINK (using the function: --tucc) and merged with the sample of children and parents.

Data from 5,081 individuals of European ancestry from the Wellcome Trust Case-Control Consortium phase-2, obtained from the 1958 British Birth Cohort and UK Blood Services collection (6), had previously been processed together with a subset of the ADHD cases (1,7). Details of imputation and quality control have been published previously (7). The sample was merged with the ADHD families, using only common autosomal SNPs that were available after all QC in both datasets.

*Polygenic risk score (PRS) calculation*

PRS were calculated for common autosomal variants in PLINK based on the following 9 large and independent neurodevelopmental/psychiatric disorder and cognitive ability discovery GWAS of primarily European ancestry: ADHD (18,378 cases and 29,113 controls) (7), anxiety disorders (31,977 cases and 82,114 controls) (8), autism spectrum disorder (ASD; 18,381 cases and 27,969 controls) (9), bipolar disorder (BD; 20,352 cases and 31,358 controls) (10), major depressive disorder (MDD; 59,851 cases and 113,154 controls) (11), schizophrenia (67,390 cases and 94,015 controls) (12), obsessive-compulsive disorder (OCD; 2,688 cases and 7,037 controls) (13), Tourette’s syndrome (TS; 4,819 cases and 9,488 controls) (14), and cognitive ability (269,867 individuals) (15). For ADHD, the GWAS meta-analysis that was used excluded the ADHD and control samples used in this study (7). PRS were re-calculated on the merged ADHD and reference population sample, but only for the discovery phenotypes with GWAS data available that did not use this population sample (i.e. ADHD, ASD, anxiety, cognitive ability, and schizophrenia). Sample sizes of the discovery GWAS were as above, except for schizophrenia, which consisted of a subset of the previous samples: 61,839 cases and 87,724controls, made available by the analysts (12).

For each discovery GWAS, we selected common (MAF>0.05) variants that overlapped with the target data and performed linkage disequilibrium (LD) clumping in PLINK (--clump-kb 500 --clump-r2 0.2) to obtain an independent set of SNPs, while retaining the most significant SNP in each LD block. For schizophrenia, we additionally excluded all variants in the extended MHC region (chromosome 6, base positions 25–35Mb) to avoid potential bias by extensive LD in this region. PRS were calculated for each individual by summing the number of alleles (weighted by the log of the odds ratio) across the set of SNPs in PLINK (with allele frequency imputation of missing data using the 1000 Genomes European ancestry reference population). We calculated PRS using 7 different p-value thresholds to select SNPs (p_T_<1, p_T_<0.5, p_T_<0.1, p_T_<0.05, p_T_<0.01, p_T_<0.001, p_T_<0.00001). The maximum number of SNPs included was as follows; for the ADHD-only sample: ADHD: 81,226, anxiety: 81,351, ASD: 81,437, cognitive ability: 74,804, MDD: 80,542, BD: 81,531, schizophrenia: 81,159, OCD: 81,447, TS: 81,662; for the merged ADHD-control sample: ADHD: 78,737, anxiety: 78,886, ASD: 79,029, cognitive ability: 72,602, schizophrenia: 78,512, OCD: 78,934, TS: 79,015. For each discovery phenotype, we then performed PCA of the correlation matrix of these 7 scores and extracted the first PC for analyses, following the PRS-PCA method, an approach that reduces overfitting and has been shown to maintain good power (16). The sign of the loadings of the PRS on the first PCs is arbitrary and therefore PRS-PCs that were negatively correlated with the raw score variables were inverted. For each of the 7 discovery phenotypes, the first PRS-PCs explained between 59.0–79.0% of the variation in the different p-value threshold PRS; this was similar (60.1–78.9%) in the merged ADHD-control sample. The PRS-PCs were standardised to be z-scores (in the ADHD-only sample) or using the mean and standard deviation of PRS in the reference population (for the merged ADHD-control sample).

PCAiR (5) was used to run PCA on an LD-pruned set of common (MAF>0.05) markers to extract the top 10 principal components to use as covariates.

*IMAGE sample*

The International Multicentre ADHD Genetics (IMAGE) data were accessed via dbGAP under approved Project #26394. This is a child sample and DSM-IV ADHD diagnosis was confirmed using the Parental Account of Children's Symptoms (PACS) interview (17). The original study exclusion criteria were autism, epilepsy, brain disorders and any genetic or medical disorder associated with externalizing behaviours that might mimic ADHD.

After basic QC exclusions (SNP missingness >0.02, individual missingness >0.02, HWE threshold p<1^-10^, heterozygosity >4SD), data were converted to genome build hg19, and imputed using the Haplotype Reference Consortium (HRC) reference panel, using the Michigan imputation server. Best guess genotypes were defined using PLINK-2, using the following QC filters: imputation info score >0.8, and MAF>0.05. Family relationships were confirmed using IBD and Mendel analyses in PLINK and Mendel errors were set to missing. Only complete parent-offspring trios were kept for analyses (N=844 complete trios). The sample is genetically diverse and primarily of Western European ancestry (from eight countries: Belgium, Germany, Ireland, Israel, the Netherlands, Spain, Switzerland, and the United Kingdom); no restrictions were made based on ancestry for the pTDT analysis.

The sample was merged with the WTCCC controls using only common autosomal SNPs that were available after all QC in both datasets and samples that were outliers on the top 5 PCs were excluded to derive a well-matched and ancestrally homogeneous sample for case-control analyses, resulting in 616 complete ADHD trios and 5,070 controls. PRS were calculated in PLINK, using the same method as above. The ADHD PRS were calculated based on an ADHD GWAS excluding the IMAGE cohort and the WTCCC controls, consisting of 14,584 cases & 22,492 controls (7).

*References*

1.	Stergiakouli E, Hamshere M, Holmans P, Langley K, Zaharieva I, Hawi Z, et al. Investigating the contribution of common genetic variants to the risk and pathogenesis of ADHD. Am J Psychiatry. 2012 Feb;169(2):186–94.

2.	Loh PR, Danecek P, Palamara PF, Fuchsberger C, Reshef YA, Finucane HK, et al. Reference-based phasing using the Haplotype Reference Consortium panel. Nat Genet. 2016;48(11):1443–8.

3.	Deelen P, Bonder MJ, Van Der Velde KJ, Westra HJ, Winder E, Hendriksen D, et al. Genotype harmonizer: Automatic strand alignment and format conversion for genotype data integration. BMC Res Notes. 2014;7(1).

4.	Das S, Forer L, Schönherr S, Sidore C, Locke AE, Kwong A, et al. Next-generation genotype imputation service and methods. Nat Genet. 2016 Oct 1;48(10):1284–7.

5.	Conomos MP, Miller MB, Thornton TA. Robust inference of population structure for ancestry prediction and correction of stratification in the presence of relatedness. Genet Epidemiol. 2015;39(4):276–93.

6.	The Wellcome Trust Case Control Consortium. Genome-wide association study of 14,000 cases of seven common diseases and 3,000 shared controls. Nature. 2007;447(7145):661–78.

7.	Demontis D, Walters RK, Martin J, Mattheisen M, Als TD, Agerbo E, et al. Discovery of the first genome-wide significant risk loci for attention deficit/hyperactivity disorder. Nat Genet. 2019 Nov 26;51:63–75.

8.	Purves KL, Coleman JRI, Meier SM, Rayner C, Davis KAS, Cheesman R, et al. A major role for common genetic variation in anxiety disorders. Mol Psychiatry. 2019 Nov 20;1–12.

9.	Grove J, Ripke S, Als TD, Mattheisen M, Walters RK, Won H, et al. Identification of common genetic risk variants for autism spectrum disorder. Nat Genet. 2019 Jan 1;51(3):431–44.

10.	Stahl EA, Breen G, Forstner AJ, McQuillin A, Ripke S, Trubetskoy V, et al. Genome-wide association study identifies 30 loci associated with bipolar disorder. Nat Genet. 2019;51(5):793–803.

11.	Wray N, Ripke S, Mattheisen M, Traylor Trzaskowski M, Byrne E, Abdellaoui A, et al. Genome-wide association analyses identify 44 risk variants and refine the genetic architecture of major depression. Nat Genet. 2018 May 1;50(5):668–81.

12.	Schizophrenia Working Group of the Psychiatric Genomics Consortium. Mapping genomic loci prioritises genes and implicates synaptic biology in schizophrenia. medRxiv. 2020 Sep 13;2020.09.12.20192922.

13.	Arnold PD, Askland KD, Barlassina C, Bellodi L, Bienvenu OJ, Black D, et al. Revealing the complex genetic architecture of obsessive-compulsive disorder using meta-analysis. Mol Psychiatry. 2018 May 1;23(5):1181–8.

14.	Yu D, Sul JH, Tsetsos F, Nawaz MS, Huang AY, Zelaya I, et al. Interrogating the Genetic Determinants of Tourette’s Syndrome and Other Tic Disorders Through Genome-Wide Association Studies. https://doi.org/101176/appi.ajp201818070857. 2019 Mar 1;176(3):217–27.

15.	Savage J, Jansen P, Stringer S, Watanabe K, Bryois J, de Leeuw C, et al. Genome-wide association meta-analysis in 269,867 individuals identifies new genetic and functional links to intelligence. Nat Genet. 2018 Jul 1;50(7):912–9.

16.	Coombes BJ, Ploner A, Bergen SE, Biernacka JM. A principal component approach to improve association testing with polygenic risk scores. Genet Epidemiol. 2020 Jul 21;44(7):gepi.22339.

17.	Neale BM, Lasky‐Su J, Anney R, Franke B, Zhou K, Maller J, et al. Genome-wide association scan of attention deficit hyperactivity disorder. Am J Med Genet Part B Neuropsychiatr Genet. 2008;3255(315):1337–44.

**Supplementary Tables**

**Table S1: Results of the polygenic transmission disequilibrium tests (pTDT)**

| Primary ADHD sample | | | | |
| --- | --- | --- | --- | --- |
| Discovery  phenotype | **mean** | **SE** | **p** | **p_FDR_** |
| ADHD | 0.30 | 0.06 | 1.3 x 10^-7^ | 5.8 x 10^-7^ |
| ANX | -0.02 | 0.06 | 0.73 | 0.77 |
| ASD | -0.07 | 0.05 | 0.17 | 0.37 |
| BD | 0.04 | 0.05 | 0.44 | 0.66 |
| COG | -0.33 | 0.05 | 2.1 x 10^-9^ | 1.8 x 10^-8^ |
| MDD | 0.02 | 0.06 | 0.75 | 0.77 |
| OCD | -0.18 | 0.05 | 1.4 x 10^-3^ | 4.3 x 10^-3^ |
| SCZ | 0.06 | 0.05 | 0.29 | 0.52 |
| TS | 0.02 | 0.06 | 0.78 | 0.77 |
| Replication ADHD sample | | | | |
| Discovery  phenotype | **mean** | **SE** | **p*** |  |
| ADHD | 0.20 | 0.03 | 2.4 x 10^-9^ |  |
| ANX | -0.14 | 0.03 | 1.8 x 10^-5^ |  |
| ASD | -0.02 | 0.03 | 0.48 |  |
| BD | -0.01 | 0.02 | 0.77 |  |
| COG | -0.06 | 0.02 | 1.2 x 10^-3^ |  |
| MDD | 0.05 | 0.03 | 0.14 |  |
| OCD | -0.08 | 0.03 | 2.6 x 10^-3^ |  |
| SCZ | -0.07 | 0.03 | 0.011 |  |
| TS | -0.01 | 0.03 | 0.62 |  |

* Replication results are presented without multiple testing correction. ADHD: attention deficit hyperactivity disorder; ANX: anxiety disorders; ASD: autism spectrum disorder; BD: bipolar disorder; COG: cognitive ability; FDR: false discovery rate multiple testing correction (primary results only); MDD: major depressive disorder; OCD: obsessive-compulsive disorder; SCZ: schizophrenia; TS: Tourette’s syndrome.

**Table S2: Comparison of polygenic risk scores in children, fathers, mothers, and non-transmitted parental alleles against the reference population sample**

|  |  | Primary sample | | |  | Replication sample | |
| --- | --- | --- | --- | --- | --- | --- | --- |
| Comparison | **Discovery phenotype** | **OR (95% CIs)** | **p** | **p_FDR_** | **OR (95% CIs)** | | **p*** |
| Children vs. Reference | **ADHD** | 1.44 (1.28-1.61) | 4.6 x 10^-10^ | 6.4 x 10^-9^ | 1.43 (1.32-1.56) | | 3.9 x 10^-17^ |
|  | **ANX** | 1.07 (0.96-1.19) | 0.24 | 0.52 | 1.13 (1.04-1.22) | | 4.3 x 10^-3^ |
|  | **ASD** | 0.96 (0.86-1.07) | 0.49 | 0.76 | 1.10 (1.01-1.20) | | 0.022 |
|  | **COG** | 0.65 (0.58-0.73) | 1.8 x 10^-13^ | 5.0 x 10^-12^ | 0.90 (0.83-0.98) | | 0.013 |
|  | **OCD** | 0.89 (0.80-0.98) | 0.020 | 0.080 | 0.98 (0.90-1.07) | | 0.71 |
|  | **SCZ** | 1.03 (0.93-1.16) | 0.55 | 0.77 | 1.02 (0.94-1.11) | | 0.57 |
|  | **TS** | 1.03 (0.93-1.15) | 0.55 | 0.77 | 1.05 (0.97-1.14) | | 0.20 |
| Non-transmitted parent alleles vs. Reference | **ADHD** | 1.01 (0.91-1.12) | 0.87 | 0.94 | 0.95 (0.87-1.03) | | 0.22 |
|  | **ANX** | 1.10 (0.98-1.23) | 0.095 | 0.30 | 1.08 (1.00-1.17) | | 0.065 |
|  | **ASD** | 1.07 (0.96-1.19) | 0.24 | 0.52 | 0.99 (0.91-1.08) | | 0.80 |
|  | **COG** | 1.01 (0.90-1.13) | 0.91 | 0.94 | 1.05 (0.97-1.15) | | 0.25 |
|  | **OCD** | 1.07 (0.96-1.19) | 0.24 | 0.52 | 1.05 (0.96-1.14) | | 0.29 |
|  | **SCZ** | 0.99 (0.89-1.10) | 0.88 | 0.94 | 0.98 (0.91-1.07) | | 0.70 |
|  | **TS** | 1.05 (0.94-1.17) | 0.36 | 0.63 | 1.01 (0.93-1.10) | | 0.84 |
| Mothers vs. Reference | **ADHD** | 1.23 (1.10-1.37) | 2.3 x 10^-4^ | 1.6 x 10^-3^ | 1.20 (1.10-1.31) | | 3.5 x 10^-5^ |
|  | **ANX** | 1.08 (0.97-1.20) | 0.17 | 0.48 | 1.20 (1.11-1.31) | | 5.0 x 10^-6^ |
|  | **ASD** | 1.04 (0.93-1.17) | 0.45 | 0.74 | 1.08 (0.99-1.17) | | 0.10 |
|  | **COG** | 0.83 (0.74-0.93) | 8.9 x 10^-4^ | 5.0 x 10^-3^ | 0.98 (0.90-1.07) | | 0.63 |
|  | **OCD** | 0.95 (0.85-1.05) | 0.32 | 0.60 | 1.00 (0.92-1.09) | | 0.95 |
|  | **SCZ** | 1.02 (0.92-1.14) | 0.67 | 0.89 | 1.06 (0.98-1.15) | | 0.16 |
|  | **TS** | 1.06 (0.95-1.19) | 0.27 | 0.54 | 1.05 (0.97-1.15) | | 0.24 |
| Fathers vs. Reference | **ADHD** | 1.18 (1.05-1.32) | 4.1 x 10^-3^ | 0.019 | 1.13 (1.03-1.23) | | 7.4 x 10^-13^ |
|  | **ANX** | 1.10 (0.99-1.21) | 0.068 | 0.24 | 1.01 (0.93-1.10) | | 0.77 |
|  | **ASD** | 1.01 (0.90-1.13) | 0.90 | 0.94 | 1.02 (0.93-1.11) | | 0.72 |
|  | **COG** | 0.79 (0.70-0.89) | 5.8 x 10^-5^ | 5.4 x 10^-4^ | 0.97 (0.89-1.05) | | 0.45 |
|  | **OCD** | 1.00 (0.89-1.11) | 0.96 | 0.96 | 1.03 (0.95-1.12) | | 0.44 |
|  | **SCZ** | 1.01 (0.90-1.12) | 0.90 | 0.94 | 0.95 (0.88-1.04) | | 0.27 |
|  | **TS** | 1.02 (0.92-1.13) | 0.73 | 0.93 | 1.01 (0.93-1.10) | | 0.79 |

* Replication results are presented without multiple testing correction.

ADHD: attention deficit hyperactivity disorder; ANX: anxiety disorders; ASD: autism spectrum disorder; COG: cognitive ability; OCD: obsessive-compulsive disorder; SCZ: schizophrenia; TS: Tourette’s syndrome.

Note: Bipolar disorder and major depressive disorder could not be examined due to inclusion of the reference population sample in the discovery genetic studies for those disorders.
